# Development and validation of a dynamic 48-hour in-hospital mortality risk stratification for COVID-19 in a UK teaching hospital: a retrospective cohort study

**DOI:** 10.1101/2021.02.15.21251150

**Authors:** Martin Wiegand, Sarah L. Cowan, Claire S. Waddington, David J. Halsall, Victoria L. Keevil, Brian D. M. Tom, Vince Taylor, Effrossyni Gkrania-Klotsas, Jacobus Preller, Robert J. B. Goudie

**Affiliations:** MRC Biostatistics Unit, School of Clinical Medicine, University of Cambridge, UK; Addenbrooke’s Hospital, Cambridge, UK; Department of Medicine, University of Cambridge, UK; Cambridge University Hospitals Trust, Cambridge, UK; Department of Medicine for the Elderly, Addenbrooke’s Hospital, Cambridge, UK; Department of Medicine, University of Cambridge; Cancer Research UK, Cambridge University Hospitals NHS Foundation Trust, Cambridge CB2 0QQ, UK; Department of Infectious Diseases, Cambridge University Hospitals, Cambridge, UK; Cambridge University Hospitals, Cambridge, UK

**Author notes:** Corresponding author: Martin Wiegand, MRC Biostatistics Unit, School of Clinical Medicine, East Forvie Building, University Forvie Site, CB2 0SR, University of Cambridge, UK. Joint last (Jacobus Preller, Robert J. B. Goudie).

**Keywords:** COVID-19, SARS-CoV-2, Mortality prediction, Dynamic prediction, Landmarking

## Abstract

**Objectives:** To develop a disease stratification model for COVID-19 that updates according to changes in a patient’s condition while in hospital to facilitate patient management and resource allocation.

**Design:** In this retrospective cohort study we adopted a landmarking approach to dynamic prediction of all cause in-hospital mortality over the next 48 hours. We accounted for informative predictor missingness, and selected predictors using penalised regression.

**Setting:** All data used in this study was obtained from a single UK teaching hospital.

**Participants:** We developed the model using 473 consecutive patients with COVID-19 presenting to a UK hospital between March 1 and September 12, 2020; and temporally validated using data on 1119 patients presenting between September 13, 2020 and March 17, 2021.

**Primary and secondary Outcomes:** The primary outcome is all-cause in-hospital mortality within 48 hours of the prediction time. We accounted for the competing risks of discharge from hospital alive and transfer to a tertiary Intensive Care Unit for extracorporeal membrane oxygenation.

**Results:** Our final model includes age, Clinical Frailty Scale score, heart rate, respiratory rate, SpO2/FiO2 ratio, white cell count, presence of acidosis (pH < 7.35) and Interleukin-6. Internal validation achieved an AUROC of 0.90 (95% CI 0.87–0.93) and temporal validation gave an AUROC of 0.86 (95% CI 0.83-0.88).

**Conclusion:** Our model incorporates both static risk factors (e.g. age) and evolving clinical and laboratory data, to provide a dynamic risk prediction model that adapts to both sudden and gradual changes in an individual patient’s clinical condition. Upon successful external validation, the model has the potential to be a powerful clinical risk assessment tool.

**Trial Registration:** The study is registered as “researchregistry5464” on the Research Registry (www.researchregistry.com).

**Article Summary:**

- Our dynamic prediction model is able to incorporate patient data as it accumulates throughout a hospital visit.
- We use the established statistical landmarking approach to dynamic prediction; account for competing risks for the primary outcome of in-hospital mortality; and the potentially-informative availability of clinical and laboratory data.
- The sample size of the first wave of patients admitted with severe COVID-19 was relatively low, due to the lower incidence in Cambridgeshire, but increased significantly during the winter months of 2020/21, providing the opportunity to temporally validate the model.
- As a single centre study, the presented model will require external validation to assess its performance in other cohorts; and also if there are significant changes in the characteristics of new variants or the management thereof.
- Our work also highlights the adaptability of the statistical landmarking framework to be used to model individual patient outcomes using densely-collected hospital data.

## Introduction

SARS-CoV-2 virus infection, the cause of COVID-19, results in a spectrum of disease ranging from asymptomatic infection through to life threatening disease requiring critical care, and even death. For patients admitted to hospital, it is essential to identify who is at risk of deterioration and death to enable timely targeted interventions (such as immune modulation and mechanical ventilation), to facilitate appropriate resource allocation and patient flow, and to inform discussions with patients and families.

Most existing disease severity prediction models for COVID-19 use only data that are available at the time of admission to hospital. Such point-of-admission models have been proposed for both mortality and composite escalation/mortality outcomes, including new and re-purposed severity and early warning scores^1-7^ and time-to-event models^8-13^.

While some markers of severity, such as sex and age can be assumed constant for the duration of the hospital visit, others, such as clinical observations and blood test results, can change markedly over the course of admission. COVID-19 is a dynamic disease in which patients can deteriorate over a short time period or suffer acute complications e.g. thromboembolism^14-15^. This may have a significant effect on a patient’s prognosis that cannot be foreseen by a point-of-admission model.

A model with the ability to adjust predictions at arbitrary time points by including updated patient information could greatly aid in clinical decision-making. Dynamic models that assimilate clinical data as it accrues may provide more accurate and clinically useful prediction of a patient’s clinical course and prognosis over the subsequent days than point-of-admission models. Predictive models that incorporate post-admission information are limited in number and scope. Some models for predicting mortality or deterioration have used information after admission, but do not continue beyond the first few days of admission^16-17^. More recent time-varying Cox models (for mortality and escalation)^18,19^ and machine learning models (for mortality)^20^ have used additional post-admission data. However, time-varying Cox models should not be used for prediction, because they require knowledge of clinical information from the future to calculate the hazard function, which is impossible in practice^21^. Furthermore, while indicating promising discrimination, these models use clinically unjustifiable or unclear methods for handling missing data and censoring, and do not account for informative missingness or consider the effect of treatments. Informative missingness describes the fact that in routinely-collected data the availability (or absence) of a result or observation may be related to the probability of the outcome. For example, a more extensive panel of investigations may be sent for patients thought more likely to benefit from escalation in care, such as transfer to an Intensive Care Unit (ICU). While often ignored, such effects can be strong in Electronic Health Record (EHR) data^22,23^.

We propose a prognostic risk stratification score for hospital patients with COVID-19, based on prediction of mortality in the subsequent 48 hours, using routinely-collected clinical data. Our model is based upon a principled statistical approach called landmarking^21,24,25^ that allows inclusion of any time-varying clinical parameters recorded prior to the time of prediction, whilst appropriately accounting for censoring and changes in the set of patients at risk. The model accounts for informative missingness and competing risks, which arise when there are two or more mutually exclusive outcomes: for example, once a patient is discharged, the risk of in-hospital mortality (during that admission) is removed. Therefore discharge is a “competing risk^”26^ when viewed from the perspective of in-hospital mortality. We account for competing risks within the landmarking framework using a recently-proposed approach^27^ that has not previously been used to model individual patient outcomes using densely-collected EHR data.

## Materials and Methods

### Study design

This is a retrospective cohort study of all patients presenting to Cambridge University Hospitals, a regional, tertiary care, university hospital in the East of England, between March 1, 2020 and March 17, 2021. This hospital is the sole admission hospital for patients in its immediate catchment population with COVID-19, and is a regional referral centre for a wide range of specialist services, which do not include extracorporeal membrane oxygenation (ECMO).

We report our findings according to the TRIPOD reporting guidelines^28^.

### Study Population

All adults (>= 18 years of age) presenting to hospital during the study period and diagnosed with COVID-19 were included. Diagnosis was based on either a positive diagnostic SARS-CoV-2 test during or up to 14 days prior to the hospital visit, or a clinical diagnosis of COVID-19 (eAppendix 1). Patients with clinically diagnosed COVID-19 (based on symptoms, and the clinical opinion of the treating clinician) were included because diagnostic testing was limited during the early stages of the pandemic^29^.

We include only the first hospital visit for each patient involving (or subsequent to) their first positive test; any re-admissions were excluded. Nosocomial infection was defined as a first positive SARS-CoV-2 test or diagnosis more than 10 days after hospital admission. Since we first train our model at 6 hours (to allow time for laboratory investigations), patients who died, were discharged or were classified as end of life within 6 hours of presentation to hospital were excluded.

All patients were treated as per detailed local guidance in use in the hospital at the time. Patients were also eligible for inclusion in relevant clinical trials running at the hospital during the study period (eAppendix 2).

### Outcomes

Throughout each patient’s hospital visit, we aim to predict all-cause in-hospital mortality during the next 48 hours, a time period that we refer to as the “prediction horizon”. We also considered two competing risks: transfer to a tertiary ICU for ECMO; and discharge from the hospital due to clinical improvement. Patients were followed up until March 19, 2021.

### Ethics

The study was approved by a UK Health Research Authority ethics committee (20/WM/0125, eAppendix 3).

### Patient and public involvement

No patients were involved in the design of this study.

### Model development

We selected a list of 59 candidate clinical parameters (eTable 1) that have been included in existing point-of-admission prediction models or were clinically judged to be likely predictors. These are divided into five categories: demographics; comorbidities; observations; laboratory tests; and treatments, interventions and level of care.

Basic patient demographics were extracted from the hospital EHR: age, sex, ethnicity, and deviation from standard ranges of Body Mass Index (BMI).

Twelve comorbidities that have previously been associated with COVID-19^30^ were identified by the presence of the corresponding ICD-10 codes entered in the EHR prior to the time at which the prediction is made (either before or during the hospital visit). eTable 2 provides the ICD-10 codes used to define each comorbidity. In addition to specific comorbidities, frailty amongst patients over 65 years old was assessed by the Clinical Frailty Scale (CFS) score^31^ (eAppendix 4).

We included the following observations that are regularly recorded in the EHR: heart rate (HR), mean arterial pressure, temperature and respiratory rate (RR). SpO2/FiO2 ratio was calculated (where SpO2 and FiO2 were available at the same timepoint) to indicate the severity of hypoxia^32-33^. SpO2 itself was not included as a potential predictor as our exploratory work suggested that, without accounting for FiO2, this largely reflected a patient’s assigned oxygen saturation targets, and therefore acted as a proxy for underlying respiratory disease (e.g. patients with chronic obstructive pulmonary disease being assigned a lower SpO2 target). Where only oxygen flow rate was available, FiO2 was estimated according to the EPIC II conversion tables^34^. PaO2/FiO2 (P/F) ratio was also included. We summarised the measurements recorded over the previous 24-hour period as follows: mean, minimum and maximum value. We also calculated the ‘median-trend’ as the difference between the median value for the last 24 hours, and the median value for the 24 hours prior to this. The Glasgow Coma Score (GCS) was extracted from the EHR; patients without a recorded GCS were assumed to have a GCS >= 12.

For each of the 31 laboratory tests we considered, we included results up to 48 hours prior to the time at which the prediction was made. Where more than one result was available, we used the most recent result. In addition, for 7 of the most frequently measured blood tests (C-reactive protein (CRP), white cell count (WCC), platelets, haemoglobin, creatinine, sodium, potassium), we included the median-trend. The neutrophil/lymphocyte and Interleukin-6/Interleukin-10 (serum IL-6/IL-10) ratios have previously been identified as prognostic, therefore we also considered these as potential predictors^9,17,35^. For blood markers where both abnormally low and abnormally high results could potentially be associated with poor prognosis (sodium and pH), we included the maximum deviation below and above the normal range in the previous 24 hours. We adjusted venous pH results by adding 0.03 to approximate arterial pH results^36^.

We included 5 indicators of treatments, interventions and levels of care. The level of care of the patient was summarised by whether the patient had been in an ICU bed in the previous 24 hours. Mechanical ventilation was defined as patients receiving invasive ventilation during the previous 24 hours, either via endotracheal tube or tracheostomy. The use of renal replacement therapy during the last 24 hours was identified from the EHR. Cardiovascular support was defined as the administration of any vasopressors or inotropes in the last 24 hours. Steroid administration has been shown to decrease the risk of death in patients with COVID-19^37-38^. We therefore include an indicator of whether the patient had received treatment dose steroids (defined as 6mg dexamethasone daily or an equivalent dose of prednisolone, hydrocortisone or methylprednisolone) during their hospital admission prior to the landmark time.

### Models

We used the landmarking approach for dynamic prediction^21,24-25^. At intervals of 24 hours (the “landmark times’’), we trained time-to-event models, using clinical parameters recorded before (or at) the landmark time as predictors. This makes it possible to include repeatedly measured clinical parameters into the prediction model, so that predictors reflect any changes in the trajectory of the patient, whilst appropriately accounting for censoring and changes in the at-risk population. If the primary outcome was recorded within the prediction horizon of a landmark time, we recorded the outcome at the relative time from landmark to event; events after the prediction horizon were censored. Patients who have had any event prior to the landmark time were excluded, since these patients were no longer at risk. The first landmark is 6 hours after presentation to allow time for clinical information to accrue, or at the point of COVID-19 diagnosis for nosocomial patients (diagnosed 10 days or more after presentation). We only used data at each landmark time from patients being actively treated for COVID-19 at that point in time. Landmark times after transition to end of life care were omitted, meaning that no predictions were made at these timepoints, although events occurring within the existing prediction horizon were still included. We used a supermodel approach in which the time-to-event model is assumed constant across landmark times^39^.

We use the Fine-Gray competing risk model to predict in-hospital death, and account for the competing risks of hospital discharge and transfer for ECMO^39,40^. A Fine-Gray model uses subdistribution hazards, which are directly related to the cumulative incidence function, by which the probability of an event of interest occuring can be estimated. While the cause-specific hazard function used by Cox models is preferable for inferring biological mechanisms, subdistribution hazard-based models are preferable for prediction^41^.

### Missing Values

We handled missing data using the missingness indicator approach since the recording in the EHR of a clinical parameter, regardless of the value, is often indicative of the treating health professional’s contemporaneous view of the patient’s condition^42,43^. Conceptually our approach, as well as estimating the “effect” of a unit increase of a particular clinical parameter (as is standard in all regression approaches), estimates the “effect” of a variable being “missing”. For each potential predictor in the model we also include a missingness indicator, which indicates that no data were recorded during the corresponding time period. This approach allows clinical parameters with an incomplete record to be included in our model and avoids the need to make the missing at random (MAR) assumption that is unlikely to hold in these data^44^. For each parameter, ultimately one of the two expressions is used for prediction for each patient at each timepoint: either a coefficient describing the relationship with the clinical parameter when it is recorded, or a fixed value if the clinical parameter is missing.

Blood tests are considered missing if the most recent measurement was collected more than 48 hours prior to the landmark time. When a blood test is repeated during the previous 48 hours, the most recent result is used. The vital signs we considered did not have missing values at any included landmark.

### Predictor Selection

To select the most predictive parameters into the model we used standard penalised variable selection, specifically Smoothly Clipped Absolute Deviations (SCAD), with the tuning parameter chosen to minimise the Bayesian Information Criterion^45^. We paired parameters together with their corresponding missingness indicator to prevent inclusion of an incompletely-recorded parameter without its missingness indicator, using the group SCAD^46^; but also allowed for the missingness indicators to be included by themselves.

The development and validation of the model has been carried out in R version 3.6^47^.

### Model assessment

Quantitative assessment of discrimination was performed using the Area Under the Receiver Opearting Characteristic (AUROC) curve, in which 0.5 indicates no discrimination and 1.0 indicates perfect discrimination. For validation of the performance of the model on the training data, in addition to the unadjusted AUROC, we also performed repeated 5-fold (split into 80% training, 20% validation data) cross-validation to account for uncertainty and over-optimism due to the complete model building process (including variable selection)^48^. We also calculated precision-recall (PR) curve and the Area Under the PR Curve (AUPRC) since it provides a clearer performance summary than AUROC when the primary outcome has low incidence, as here^49^. We assessed clinical benefit visually via the number needed to evaluate (NNE), defined as 1/positive predictive value (1/PPV), against the sensitivity. We also calculated the net benefit curve^50^.

We assessed calibration visually using a calibration plot of predicted risk against observed mortality rate. We also quantitatively assessed the calibration slope and calibration-in-the-large^51^.

### Sensitivity analyses

To assess whether the model is unduly influenced by patients with long hospitalisations we re-trained the model using only each patient’s first 28 landmark times (spanning 28 days). We also assessed the sensitivity of our model assessment by stratifying by whether COVID-19 was confirmed by a positive SARS-CoV-2 diagnostic test, and according to whether patients had received at least a single COVID-19 vaccination dose (Oxford-AstraZeneca Covishield, Pfizer-BionTech Comirnaty or Moderna Spikevax).

## Results

### Development of prediction model

We developed the model using data from Wave 1 (March 1, 2020 to September 12, 2020), with the end date chosen since only a single patient remained in hospital with COVID-19 on this date. 519 patients presented to hospital with COVID-19 during Wave 1, of whom 46 were excluded due to discharge (34), death (2) or transition to end of life care (10) prior to the first landmark time (i.e. within 6 hours of presentation). The baseline characteristics of the 473 patients included in the development of the model are shown in Table 1.

**Table 1:** Cohort demographics and clinical features in Wave 1 and Wave 2.

| **Characteristic** | **Wave 1 (training data set)** | **Wave 2 (validation data set)** |
| --- | --- | --- |
| Admission dates | Mar 1, 2020 – Sep 12, 2020 | Sep 13, 2020 – Mar 17, 2021 |
| Number of patients | 473 | 1119 |
| Female, n (%) | 196 (41.4%) | 579 (48.3%) |
| Age at admission, median [IQR], years | 69 [55, 81] | 65 [49, 79] |
| Admission BMI, median [IQR], kg/m <sup>a</sup> | 25.7 [22.1, 29.9] | 27.35 [23.5, 32.1] |
| Clinical Frailty Score at admission for over 65 year olds, median [IQR] | 5 [3, 6] | 5 [4, 6] |
| Nosocomial infection, n (%) | 36 (7.6%) | 116 (9.7%) |
| Length of stay, median [IQR], days | 10.8 [3.9, 19.8] | 8.0 [2.95,-18.5] <sup>b</sup> |
| **Ethnicity, number (%)** |  |  |
| White | 354 (74.8%) | 792 (66.1%) |
| Asian | 24 (5.1%) | 61 (5.1%) |
| Black | 10 (2.1%) | 13 (1.1%) |
| Other | 11 (2.3%) | 39 (3.3%) |
| Prefer not to say/ Not recorded | 74 (15.6%) | 294 (24.5%)) |
| **Outcomes, number (%)** |  |  |
| Deceased in-hospital <sup>c</sup> | 99 (20.9%) | 145 (12.1%) |
| Transferred for ECMO | 5 (1.1%) | 1 (0.1%) (0%) |
| Discharged alive | 369 (78.0%) | 967 (80.7%) |
| Remain in hospital on Mar 19, 2021 | 0 (0%) | 86 (7.2%) |
| **Support / treatments received during hospital stay, number (%)** |  |  |
| ICU admission | 103 (21.8%) | 238 (19.8%) |
| Invasive mechanical ventilation | 82 (17.3%) | 172 (14.3%) |
| Non-invasive ventilation (CPAP or Bi-PAP) | 37 (7.8%) | 144 (12.9%) |
| Cardiovascular support | 86 (18.2%) | 170 (14.2%) |
| Renal replacement therapy | 32 (6.8%) | 39 (3.3%) |
| Steroid treatment | 120 (25.4%) | 619 (55.3%) |
| **Admission observations <sup>a</sup>, median [IQR]** |  |  |
| Mean arterial pressure, mmHg | 86 [76, 96] | 88 [78, 98] |
| Heart rate, beats/min | 83 [72, 93] | 81 [72, 91] |
| Temperature, degrees celsius | 37.2 [36.7, 37.8] | 36.9 [36.5, 37.4] |
| Respiratory rate, breaths/min | 19 [17, 22] | 18 [17, 20] |
| Oxygen saturation (SpO2), % | 96 [94, 97] | 95 [94, 97] |
| SpO2/FiO2 ratio | 448 [337, 457] | 452 [400, 462] |
| **Admission blood results <sup>a</sup>, median [IQR]** |  |  |
| Urea, mmol/L | 7.8 [5.0, 11.8] | 7.1 [5.1, 10.5] |
| Creatinine, μmol/L | 75 [61, 106] | 70 [56, 93] |
| Sodium, mmol/L | 137 [134, 140] | 137 [135, 140] |
| CRP, mg/L | 87 [39, 178] | 55 [26, 109] |
| WCC, 10<sup>9</sup>/L | 6.9 [4.9, 9.3] | 6.8 [5.0, 9.8] |
| Neutrophils, 10<sup>9</sup>/L | 5.4 [3.6, 7.8] | 5.3 [3.5, 7.9] |
| Lymphocytes, 10<sup>9</sup>/L | 0.8 [0.5, 1.2] | 0.8 [0.5, 1.2] |
| D-dimer, ng/ml | 334 [177, 682] | 292 [167, 641] |
| Troponin, ng/L | 20.0 [8.3, 63.6] | 11.0 [4.0, 39.5] |
| pH | 7.43 [7.37, 7.46] | 7.44 [7.39, 7.46] |
| IL-6, pg/ml | 15.1 [5.3, 40.2] | 11.6 [4.0, 29.8] |

<sup>a</sup> First after positive SARS-CoV-2 test result for hospital-acquired COVID-19 cases
<sup>b</sup> For Wave 2, includes stays completed by March 19, 2021 only. Note that because follow-up continued until March 19 2021, the outcome during the 48-hour prediction horizon was known for all landmark times up to March 17 2021, and so patients remaining in hospital at the study end-date could be included in the validation.
<sup>c</sup> Total in-hospital mortality including those patients who were classified as “end of life” prior to death.

In total we included 6846 landmark times for training the model, with a median of 9 (IQR 3–17) landmark times per patient. In the 48-hour prediction horizon following these landmark times, there were 119 in-hospital death events (1.7% of landmarks), 658 hospital discharge events (9.6%) and 10 transfers for ECMO (0.1%). Note that, since landmarks occur every 24 hour and the prediction horizon is 48 hours, patient events will usually occur within the prediction horizon of two adjacent landmark times. eTable 1 reports summary statistics, missingness and the number of measurements available per landmark time for each predictor. No patients were excluded due to missing data.

### Model results

Our proposed model (Table 2 and eTable 3) for 48-hour in-hospital mortality includes age, Clinical Frailty Scale (CFS) score^52^, heart rate (HR), respiratory rate (RR), oxygen saturation/fraction of inspired oxygen (SpO2/FiO2) ratio^32,33^, white cell count (WCC), acidosis (pH < 7.35) and Interleukin-6 (IL-6). The mortality probability can be calculated using the calculator at http://shiny.mrc-bsu.cam.ac.uk/apps/covid19mortalityrisk/; see eAppendix 5 for details.

**Table 2.** Final model coefficients

| Predictor | Coefficients when recorded | Coefficients if unrecorded<sup>1</sup> |
| --- | --- | --- |
| Age <75 years, at admission | -0.516 | – |
| Age <80 years, at admission | -0.245 | – |
| Clinical Frailty Score, at admission | 0.0678 | 0.0513 |
| Heart rate, beats/min, mean during last 24h | 0.00282 | – |
| Respiratory rate, breaths/min, minimum during last 24h | 0.102 | – |
| SpO2/FiO2 ratio, minimum during last 24h | -0.0116 | – |
| WCC, 10<sup>9</sup>/L, most recent measurement during last 48h | 0.00651 | -0.0015 |
| Acidosis, 7.35 - (lowest pH during last 24h), or 0 if all above 7.35 | 3.18 | 0.22 |
| IL-6, pg/ml, most recent measurement during last 48h | 0.000166 | -0.0218 |

1. If the predictor value is not recorded, the fixed value in this column is used, and the coefficient corresponding to the predictor value is ignored.

The model trained on only data from the first 28 landmarks for each patient closely resembled the model in Table 2, although the Clinical Frailty Score and IL-6 were not selected (eTable 4).

### Internal performance assessment

Figure 1 shows the internal performance metrics for the model in Table 2 (using the training data). The unadjusted internal area under receiver-operating characteristic curve (AUROC) was 0.90 (95% CI 0.87–0.93) and the median cross-validation AUROC was 0.87, both indicating good discrimination (Figure 1A). The PR curve (Figure 1B) also showed good discrimination, with an AUPRC of 0.31, in a population with 48 hour in-hospital mortality of 0.017 (1.7%), and the NNE <10 for sensitivity less than 0.75 (Figure 1C). Figure 1D shows the calibration plot. The calibration intercept was -0.02 (95% CI -0.22– 0.17) indicating that the mean predicted probabilities matched the mean observed mortality, while the calibration slope was 1.16 (95% CI 1.02–1.31) suggesting that the observed mortality in high predicted risk patients slightly exceeded the predicted mortality risk. The net benefit curve for risk stratification by the proposed model is clearly higher than for the two non-model alternatives of classifying either everyone, or no-one as high risk patients, indicating the clinical utility of the dynamic model (eFigure 3).

**Figure 1.**
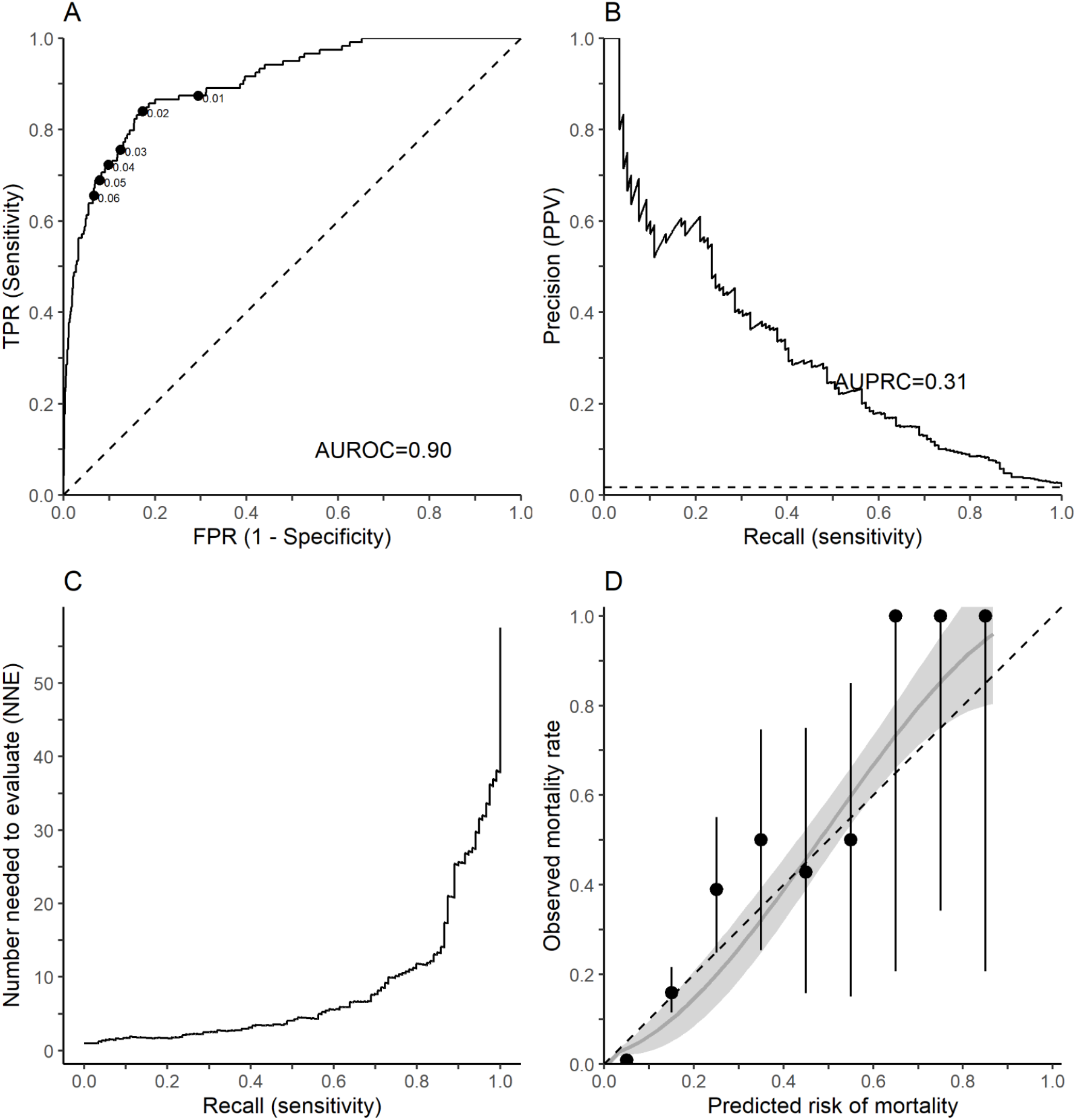
Performance metrics for in-hospital mortality in the training dataset. (A) Receiver operator characteristic plot, with labels indicating the corresponding threshold and the dashed line indicating the line of no discrimination. (B) Precision-recall plot, with the 2.8% observed incidence indicated by the dashed line. (C) Number needed to evaluate against sensitivity. (D) Calibration plot (with 95% CI), by tenths of predicted risk and a LOESS interpolation (grey), with the dashed line indicating perfect calibration.

### Temporal validation of prediction model

We assessed the performance of the model by applying it to held-out data corresponding to admissions during Wave 2 (September 13, 2020 and March 171, 2021). 1119 patients presented to the study hospital during this period. In total we tested the model using 12981 landmark times, with a median of 6 (2-14) landmark times per patient. In the 48-hour prediction horizon following these landmark times, there were 253 in-hospital death events (1.9% of landmarks), 1615 hospital discharge events (12.4%) and 2 transfers for ECMO (0.015%). 47 landmark times were omitted due to missing vital sign data. Characteristics are summarised in Table 1. Of note, compared to Wave 1, patients presenting in Wave 2 were slightly younger and more likely to be female, evidenced by a more balanced data set.

Figure 2 shows the temporal validation performance metrics, obtained by applying the trained model (Table 2) to the Wave 2 patients. The receiver-operating characteristic (ROC) curve (Figure 2A) shows the model continues to discriminate well, with AUROC 0.86 (95% CI 0.83-0.88). The PR curve (Figure 2B) shows that PPV was consistently well above the 48-hour in-hospital mortality incidence of 0.019 (1.9%) in the Wave 2 cohort across all sensitivities, with AUPRC 0.15, and NNE < 10 for sensitivities between 0.02 and 0.63 (Figure 2C). Figure 2D shows the calibration plot, which shows a tendency of the model to underpredict risk in the higher risk patients: calibration-in-the-large was 0.35 (95% CI 0.21-0.47)), suggesting the mean of the predicted probabilities was lower than the mean observed mortality, and calibration slope was 0.90 (95% CI 0.82-0.99), indicating that the spread of predicted risk corresponds reasonably well with the spread of observed mortality. The calibration plot shows a good correspondence between observed mortality rate and predicted mortality risk for the lower risk patients (<0.4), but due to the low incidence of mortality events among the landmarks corresponds less well for the higher risk patients. This is evidenced by the considerable confidence intervals. The net benefit curves for the proposed model surpasses both alternatives of classifying everyone, and no-one as high risk patients (eFigure 4).

**Figure 2.**
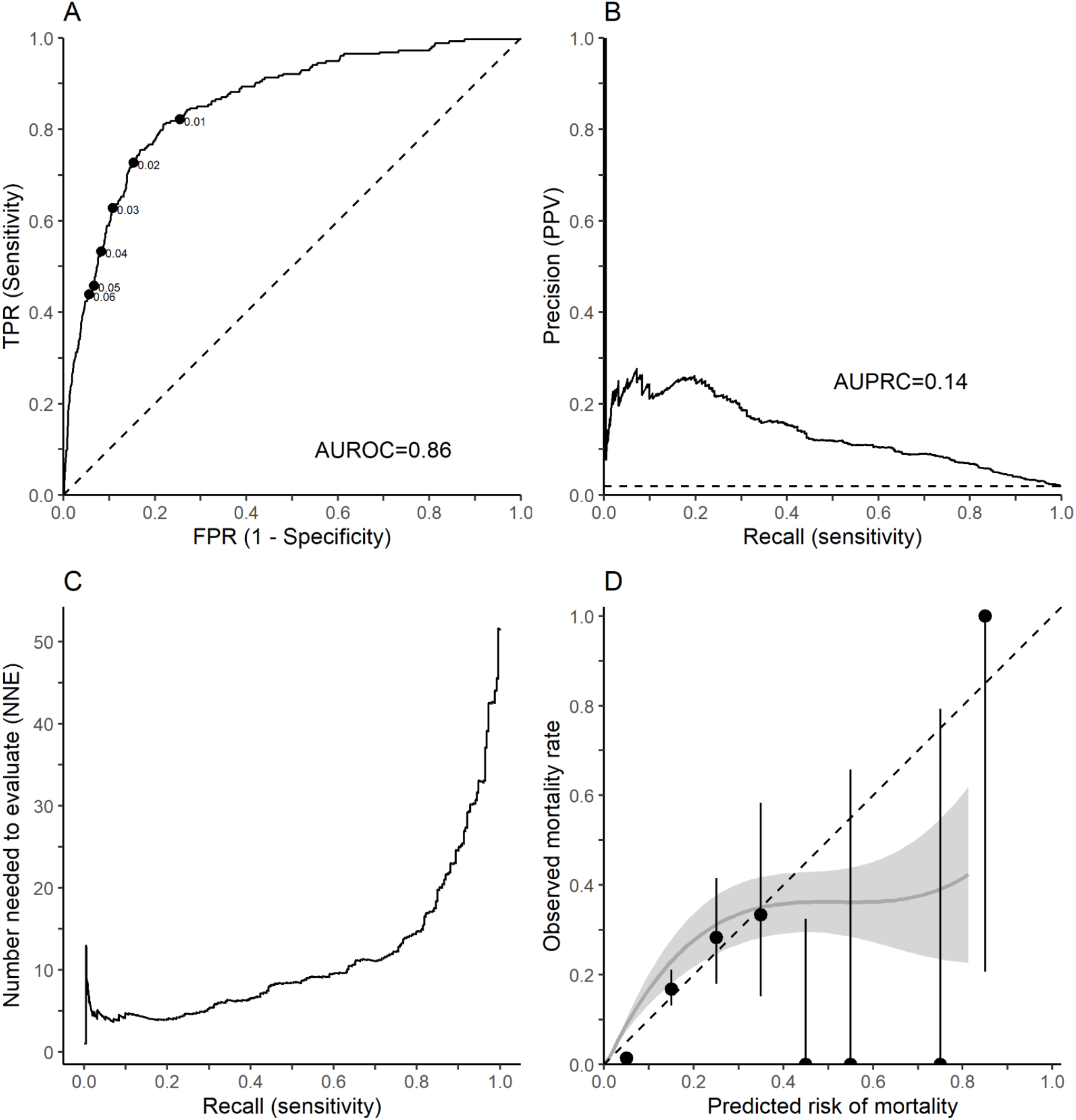
Performance metrics for in-hospital mortality in the validation dataset. (A) Receiver operator characteristic plot, with labels indicating the corresponding threshold and the dashed line indicating the line of no discrimination. (B) Precision-recall plot, with the 3.1% observed incidence indicated by the dashed line. (C) Number needed to evaluate against sensitivity. (D) Calibration plot (with 95% CI), by tenths of predicted risk and a LOESS interpolation (grey), with the dashed line indicating perfect calibration.

### Sensitivity analyses

We did not find evidence that the discrimination of the model was affected by the presence or absence of positive diagnostic SARS-CoV-2 results (rather than solely a clinical diagnosis). In patients with a positive SARS-CoV-2 test the AUROC was 0.90 (95% CI 0.87-0.93) in the training dataset and 0.85 (95% CI 0.83-0.88) in the validation dataset; whereas for patients with only a clinical diagnosis the AUROC was 0.94 (95% CI 0.88-1.00) in the training dataset and 0.88 (95% CI 0.79-0.99) in the validation dataset.

A small number of patients (65 patients, for whom 874 landmarks were available) in our validation dataset had received a COVID-19 vaccine: the AUROC of 0.88 [0.71, 0.99] for these patients suggested good discrimination and is consistent with the unvaccinated patients (AUROC 0.85 [0.83, 0.88]).

## Discussion

SARS-CoV-2 causes a wide spectrum of disease that can evolve over time, and may necessitate critical care management and even result in death. In light of the threat of further waves of coronavirus infections there is still a pressing clinical need to be able to anticipate disease severity and the trajectory of illness to facilitate patient management and resource allocation. The model described herein incorporates both static admission risk factors (age and CFS) and evolving clinical and laboratory data, providing a dynamic 48-hour risk prediction model that can adapt to both sudden and gradual changes in an individual patient’s clinical condition. The data used in the model were routinely collected demographic and clinical data from during the patient’s hospitalisation, automatically extracted from patient EHRs. As such, this model could be readily incorporated into routine clinical care.

Several methodological aspects of our approach strengthened our model. Firstly, we accounted for competing risks, whereby the outcome (risk) of interest (in this case in-hospital mortality) can only happen whilst the patient is in hospital, and therefore the outcome of interest is ‘competing’ against the risk of transfer to another hospital and/or discharge from hospital. Allowance for this is important in predictive modeling^53^. Secondly, allowance was made for the potential of the availability of observations and investigations to in itself be a reflection of disease severity. While multiple imputation is often used in clinical prediction models because it gives unbiased estimates under the MAR assumption, it is unlikely that this assumption holds in the routinely-collected EHR data that we use^42^. The missing indicator method that we adopted does not rely on the MAR assumption and can improve predictive performance in EHR data^42-44^. Furthermore, we validated our model using data from different waves. As each wave included COVID-19 variants of different infectiousness that are potentially associated with different morbidity and mortality risks, the fact that our model holds across waves further attests to the fact that our selected parameters are useful for prognostication in different clinical scenarios. Finally, we did not seek to make prognostic predictions for patients after clinicians have identified them as entering the last few hours or days of life. Since observations and investigations are often discontinued at the end of life, including these time periods would distort the model due to extreme missingness (in our data, no vital sign observations were recorded on 43% of days during end-of-life care, compared to 0% of days during active treatment). In addition, predicting end-of-life after it is clinically apparent would have little clinical utility.

Several predictors of disease severity included in our model have also been identified by point-of-admission severity models, and in epidemiological studies of risk factors for severe disease. Increasing age is widely recognised as being the strongest predictor of poor outcome from COVID-19^3,11,30^. Frailty has similarly been shown to be a strong independent predictor of mortality in hospitalised older adults^55^, including those with COVID-19^52,55^.

Respiratory compromise is a common reason for hospital admission and markers of respiratory function, including respiratory rate^3,4,12^, SpO2^3^, oxygen requirement^3^ and SpO2/FiO2 ratio have been included in previous point-of-admission models. The SpO2/FiO2 ratio, as selected by our model, allows a fully quantitative rather than dichotomous measure of the need for additional oxygen, as well as allowing for the confounding effect of variation in the target oxygen saturations in different patient groups.

Acidosis frequently complicates respiratory, renal and advanced circulatory failure and has previously been noted as a marker of disease severity in COVID-19^59^. The separate inclusion of the severity of acidosis and alkalosis in our set of candidate predictors allowed for pH changes in either direction to be accounted for and avoided, for example, a minor negative effect of alkalosis from masking a more major effect of acidosis.

Our model selected two markers of infection and inflammation: WCC and IL-6. This is consistent with other findings^11,56-57^. IL-6 was included in the routine COVID-19 panel of blood tests at the study hospital but we recognise that this may be less commonly requested in other hospitals. To assess whether C-reactive protein (CRP) could serve as a proxy for IL-6 in our model when it is not available, we refitted the model with CRP in place of IL-6 (eTables 5-6). The AUROC was slightly lower on both training (0.89, 95% CI 0.85–0.93) and validation (0.84, 95% CI 0.81–0.87) data, yielding a slightly weaker but potentially more broadly applicable model. The preference of the model for IL-6 over CRP may reflect the fact that IL-6 is responsible for the production of CRP and, as such, is an earlier and more dynamic marker of the inflammatory response^58^.

To assess whether a simpler model could perform similarly we considered removing the laboratory tests from our model (eTable 7-8). The resulting model provided slightly less good discrimination with an AUROC of 0.88 (95% CI 0.85-0.90) in the training data and an AUROC of 0.85 (95% CI 0.82-0.87) in the validation data.

To assess the feasibility of our approach with an extended time horizon and the stability of the predictors selected, we refitted the model with a 72 hour prediction horizon. The resulting model matched the previous AUROC in the training data of 0.90 and the AUROC of 0.85 in the validation data set. The full performance metrics are available in eFigure 1 and eFigure 2, and the model coefficients in eTable 9.

There are several limitations to our study. We chose to include only laboratory results up to 48 hours and vital signs up to 24 hours before the landmark time; exploiting older data might improve the predictive ability of our model, at the expense of complexity and real-world utility. Our data were gathered from a single centre, and therefore the generalisability of our findings to other centres and populations are pending external validation. Further, our model was generated from a relatively modest sample size due to the relatively low prevalence of COVID-19 patients in the catchment population of the hospital, particularly during the early months of the pandemic. One advantage of using this single dataset from a large, tertiary hospital was that the hospital never became overwhelmed with patients, and therefore it is considered that patients received care according to what was considered clinically appropriate rather than what resources permitted. Finally, while it is encouraging that the model continued to perform well in the Wave 2 validation data, changes in clinical care of patients (notably use of steroids and IL-6 inhibitors) and the dominant virus strains (including the Delta variant that emerged in the UK whilst this manuscript was under review) may influence the clinical picture of the disease, its severity and the risk factors for disease. The model will therefore likely need to be updated as the pandemic evolves, but the utilisation of routinely available data in this model makes this straightforward.

## Data Availability

The de-identified data that support the findings of this study are available from Cambridge University Hospitals but restrictions apply to the availability of these data, which were used under license for the current study, and so are not publicly available. Data are however available from the authors upon reasonable request with permission of Cambridge University Hospitals.

## Contributors

### Conception

JP, RJBG, SLC, EGK, CW and MW conceived the research project and designed the study.

### Data collection

VT, MW, SLC and RJBG extracted and curated the dataset. VLK and DJH collected data.

### Analysis tools & Data interpretation

MW, RJBG and BDMT analysed and contributed to the selection and creation of analysis tools. JP, CSW, EGK, DJH, VLK and BDMT interpreted the data and results.

### Implementation

MW and RJBG performed the implementation and analysis of the proposed method.

### Draft writing

MW, CSW, EGK, SLC and RJBG wrote the initial draft. All authors contributed to substantively revising the article for important intellectual content.

## Approval of final submission

All authors approved of the final version of this script to be published, and are accountable for the work presented.

## Acknowledgements

We thank Christopher Osuafor, Catriona Davidson, Alistair Mackett, Marie Goujon and Lelane Van Der Poel for assisting with the frailty sub-study.

