## Supplemental material for "Development and validation of a dynamic 48-hour in-hospital mortality risk stratification for COVID-19 in a UK teaching hospital: a retrospective cohort study"

**Supplementary Materials**

**eTable 1**. Complete list of clinical parameters that are candidate predictors, summary statistics and missingness for the training dataset.

| Marker | Unit | Summary measure/coding | Summary across landmark times [a] | Missingness across landmark times [b] |
|---|---|---|---|---|
| **Demographics** | | | | |
| Age at admission | Years | | 69 [55,81] | – (0%) |
| | | <45 | 517(7.6%) | – |
| | | <50 | 853 (12.5%) | – |
| | | <55 | 1386 (20.2%) | – |
| | | <60 | 2164 (31.6%) | – |
| | | <65 | 2779 (40.6%) | – |
| | | <70 | 3371 (49.2%) | – |
| | | <75 | 4284 (62.6%) | – |
| | | <80 | 5236 (76.5%) | – |
| | | <85 | 5857 (85.6%) | – |
| | | <90 | 6315 (92.3%) | – |
| | | <95 | 6670 (97.4%) | – |
| Sex | Female/ Male | | 41.4% / 58.6% | – (0%) |
| Ethnicity | | Not included | | |
| White | yes/no | White British/ White Irish/ Other white background | 5215 (76.2%) | – (16.3%) |
| Asian | yes/no | Asian Indian/ Asian Pakistani/ Asian Bangladeshi/ Other Chinese/ Other Asian background | 225 (3.3%) | – (16.3%) |

| | | | | |
|---|---|---|---|---|
| Black | yes/no | Black Carribean/ Black African/ Other Black background | 155 (2.3%) | – (16.3%) |
| Other | yes/no | Other ethnic group/ Mixed white and black carribean/ Mixed white and black African/ Mixed White and Asian/ Other mixed background | 137 (2.0%) | – (16.3%) |
| Body Mass Index | kg/m$^2$ | Not included | 27.3 [22.5, 30.3] | – (6.1%) |
| Underweight | kg/m$^2$ | 18.5 - (Most recent BMI), or 0 if most recent BMI above 18.5 | 93.5% = 0<br>After excluding zero: 1.4 [0.5, 1.9] | – (6.1%) |
| Overweight | kg/m$^2$ | (Most recent BMI) - 25, or 0 if most recent BMI below 25 | 38% = 0<br>After excluding zero: 4.4 [2.3, 9.1] | – (6.1%) |
| Clinical Frailty Scale [c] | | Value | 5 [3, 6] | – (54.5%) |
| **Comorbidities** | | | | |
| Asthma | yes/no | Documented history of | 780 (11.4%) | - |
| Dementia | yes/no | Documented history of | 353 (5.2%) | - |
| Diabetes | yes/no | Documented history of | 1233 (18.0%) | - |
| Chronic heart | yes/no | Documented history of | 1260 (18.4%) | - |

| | | | | |
|---|---|---|---|---|
| disease | | | | |
| Hypertension | yes/no | Documented history of | 2193 (32.0%) | - |
| Immunocompromised | yes/no | Documented history of | 80 (1.1%) | - |
| Chronic liver disease | yes/no | Documented history of | 640 (9.4%) | - |
| Non-haematological malignancy | yes/no | Documented history of | 576 (8.4%) | - |
| Haematological malignancy | yes/no | Documented history of | 284 (4.1%) | - |
| Chronic kidney disease | yes/no | Documented history of | 502 (7.3%) | - |
| Respiratory disease | yes/no | Documented history of | 833 (12.2%) | - |
| Stroke | yes/no | Documented history of | 220 (3.2%) | - |
| **Observations** | | | | |
| Heart rate (HR) | Beats/min | 24h mean | 83 [73, 93] | 17.3 (0.0%) |
| | | 24h min | 72 [63, 82] | |
| | | 24h max | 94 [83, 106] | |
| | | Trend | 0 [-4.5, 4] | – (6.3%) |
| Mean arterial pressure | mmHg | 24h mean | 86 [79, 94] | 17.1 (0.0%) |
| | | 24h min | 74 [66, 83] | |
| | | 24h max | 100 [91, 109] | |
| | | Trend | 0.0 [-0.4, 0.4] | – (6.3%) |
| Temperature | Degrees | 24h mean | 37.0 [36.7, 37.3] | 9.3 (0.0%) |

| | | | | |
|---|---|---|---|---|
| | Celsius | 24h min | 36.4 [36.1, 36.7] | |
| | | 24h max | 37.5 [37.1, 38.1] | |
| | | Trend | 0 [-0.3, 0.2] | – (6.3%) |
| Respiratory Rate (RR) | Breaths/min | 24h mean | 18.5 [17, 21] | 18.8 (0.0%) |
| | | 24h min | 16 [15, 18] | |
| | | 24h max | 20 [19, 26] | |
| | | Trend | 0 [-1,1] | – (6.3%) |
| SpO2/FiO2 ratio | | 24h mean | 431 [325, 456] | 12.8 (0.0%) |
| | | 24h min | 392 [250, 448] | |
| | | 24h max | 457 [443, 467] | |
| | | Trend | 0 [-7.0, 7.4] | – (6.3%) |
| P/F ratio | mmHg | 24h mean | 184 [136, 250] | 1.5 (77.9%) |
| | | 24h min | 140 [98, 201] | |
| | | 24h max | 229 [171, 310] | |
| | | Trend | 2 [-18, 24] | – (80.3%) |
| Glasgow coma scale (GCS) | Lowest GCS in the last 24h | <9 | 33.8% | – |
| | | <12 | 47.0% | – |
| **Laboratory tests** | | | | |
| Urea | mmol/L | Most recent measurement during last 48h | 8.8 [5.6, 14.1] | 0.9 (38.0%) |
| Creatinine | $\mu$mol/L | Most recent measurement during last 48h | 70 [52, 106] | 0.9 (25.8%) |
| | | Trend | -1 [-7, 4] | – (43.4%) |
| Sodium | mmol/L | Not included | 138.6 [135.5, 142] | 2.7 (24.8% |
| | | Trend | 0.0 [-1.0, 1.5] | – (41.9%) |
| Hyponatraemia | Na < 135 mmol/L | 135 - (lowest sodium during last 24h), or 0 if all above 135 | 82% = 0<br>After excluding zero: 2 [1, 4] | 2.7 (24.8%) |

| | | | | |
|---|---|---|---|---|
| Hypernatraemia | Na > 145 mmol/L | (highest sodium during last 24h) - 145, or 0 if all below 145 | 85.2% = 0 After excluding zero: 3.8 [2, 6.2] | |
| Potassium | mmol/L | Most recent measurement during last 24h | 4.1 [3.7, 4.4] | 2.7 (25.0%) |
| | | Trend | 0 [-0.2, 0.2] | – (42.1%) |
| Albumin | g/L | Most recent measurement during last 48h | 24 [20, 28] | 1.2 (33.6%) |
| Alanine Transaminase (ALT) | U/L | Most recent measurement during last 48h | 36 [22, 61] | 0.7 (37.2%) |
| Alkaline phosphatase (ALP) | U/L | Most recent measurement during last 48h | 100 [73, 149] | 2.3 (19.8%) |
| Bilirubin | $\mu$mol/L | Most recent measurement during last 48h | 8 [5, 13] | 0.7 (37.4%) |
| Lactate dehydrogenase (LDH) | U/L | Most recent measurement during last 48h | 335 [261, 436] | 0.2 (80.5%) |
| C-reactive protein (CRP) | mg/L | Most recent measurement during last 48h | 56 [22, 131] | 1.6 (12.8%) |
| | | Trend | 0 [-0.2, 0.3] | – (45.9%) |
| Procalcitonin (PCT) | ng/ml | Most recent measurement during last 48h | 0.24 [0.08, 0.83] | 0.2 (84.4%) |
| Ferritin | $\mu$g/L | Most recent | 726 [336, 1427] | 0.3 (76.6%) |

| | | | | |
|---|---|---|---|---|
| | | measurement during last 48h | | |
| Haemoglobin | g/L | Most recent measurement during last 48h | 104 [89, 122] | 5.4 (12.4%) |
| | | Trend | -1 [-5, 3] | – (43.9%) |
| White cell count (WCC) | $10^9$/L | Most recent measurement during last 48h | 7.9 [5.7, 10.6] | 1.6 (12.7%) |
| | | Trend | 0 [-1, 1] | – (46.1%) |
| Neutrophils | $10^9$/L | Most recent measurement during last 48h | 5.7 [3.9, 8.2] | 1.6 (13.4%) |
| Lymphocytes | $10^9$/L | Most recent measurement during last 48h | 1.1 [0.7, 1.5] | 1.6 (13.4%) |
| Neutrophil-Lymphocytes ratio | Ratio | Most recent measurement during last 48h | 5.4 [3.2, 9.5] | 1.6 (13.5%) |
| Eosinophils | $10^9$/L | Most recent measurement during last 48h | 0.1 [0.02, 0.28] | 1.6 (14.1%) |
| Monocytes | $10^9$/L | Most recent measurement during last 48h | 0.45 [0.3, 0.64] | 1.6 (13.5%) |
| Platelets | $10^9$/L | Most recent measurement during last 48h | 280 [193, 387] | 1.6 (12.8%) |
| | | Trend | 3 [-17, 26] | – (46.3%) |
| Red cell distribution width (RDW) | % | Most recent measurement during last 48h | 15.1 [14, 16.4] | 1.6 (13.3%) |
| Prothrombin Time | sec | Most recent | 13.3 [12.5, 14.5] | 0.84 (52.6%) |

| | | | | |
|---|---|---|---|---|
| | | measurement during last 48h | | |
| Activated partial thromboplastin time (APTT) | sec | Most recent measurement during last 48h | 32.3 [29.6, 35.3] | 0.84 (53.8%) |
| D-Dimer | ng/ml | Most recent measurement during last 48h | 552 [284, 1677] | 0.4 (71.9%) |
| Troponin | ng/L | Most recent measurement during last 48h | 17 [5.6, 48.7] | 0.3 (80.5%) |
| Interferon Gamma (IG) | pg/ml | Most recent measurement during last 48h | 0.9 [0.9, 2.5] | 0.2 (84.2%) |
| TNF-Alpha (TNFA) | pg/ml | Most recent measurement during last 48h | 12.3 [8.3, 18.2] | 0.2 (84.2%) |
| Interleukin-1 beta (IL-1) | pg/ml | Most recent measurement during last 48h | 0.5 [0.3, 0.9] | 0.2 (84.2%) |
| Interleukin-6 (IL-6) | pg/ml | Most recent measurement during last 48h | 13.6 [4.7, 31.9] | 0.3 (84.0%) |
| Interleukin-10 (IL-10) | pg/ml | Most recent measurement during last 48h | 1.88 [0.7, 4.4] | 0.2 (84.2%) |
| Interleukin ratio (IL-ratio, IL6/IL10) | Ratio | Most recent measurement during last 48h | 7.6 [2.9, 20.1] | 0.2 (84.3%) |
| Lactate | mmol/L | Most recent measurement during last 48h | 1.3 [1.0, 1.7] | 3.7 (60.4%) |

| | | | | |
|---|---|---|---|---|
| pH – arterial or (venous + 0.03) | | Not included | 7.41 [7.36, 7.44] | 1.9 (66.4%) |
| Acidosis | pH-value < 7.35 | 7.35 - (lowest pH during last 24h), or 0 if all above 7.35 | 67.1% = 0<br>After excluding zero:<br>0.06 [0.028,0.107] | 1.9 (66.4%) |
| Alkalosis | pH-value > 7.45 | (Highest pH during last 24h) - 7.45, or 0 if all below 7.45 | 66.8%= 0<br>After excluding zero:<br>0.024 [0.011, 0.039] | |
| **Treatments, interventions and level of care** | | | | |
| Visited ICU | yes/no | During last 24h | 1789 (26.1%) | – |
| Mechanically ventilated | yes/no | During last 24h | 1420 (20.7%) | – |
| Cardiovascular support | yes/no | During last 24h | 705 (10.3%) | – |
| Renal replacement therapy | yes/no | During last 24h | 348 (5.1%) | – |
| Steroids (oral or intravenous; dexamethasone, hydrocortisone, prednisolone, methylprednisolone, in treatment dosage) | yes/no | Ever over a 24h prior to the landmark | 2550 (37.2%) | – |

[a] For yes/no items shown as number (%) across landmark times; for quantitative items shown as median [IQR] across landmark times

[b] Shown as mean number of measurements per landmark (% landmarks with no measurement)

[c] For 45 patients for whom no CFS score had been recorded by the treating team, a consultant or specialist registrar in Geriatric Medicine reviewed the clinical records and assigned a CFS score using only information recorded at the time of admission.

**eTable 2.** ICD-10 codes used to identify comorbidities.

| Diagnosis | ICD-10 codes | Description |
|---|---|---|
| **Hypertension** | I10 | Essential hypertension |
| | I11 | Hypertensive heart disease |
| | I12 | Hypertensive renal disease |
| | I13 | Hypertensive heart and renal disease |
| | I15 | Secondary hypertension |
| **Diabetes** | E10 | Type 1 diabetes mellitus |
| | E11 | Type 2 diabetes mellitus |
| | E12 | Malnutrition-related diabetes mellitus |
| | E13 | Other specified diabetes mellitus |
| | E14 | Other unspecified diabetes mellitus |
| **Chronic liver disease** | K70 | Alcoholic liver disease |
| | K71 | Toxic liver disease |
| | K72 | Hepatic failure, not elsewhere classified |
| | K73 | Chronic hepatitis, not elsewhere classified |
| | K74 | Fibrosis and cirrhosis of the liver |
| | K75 | Other inflammatory diseases of the liver |
| | K76 | Other diseases of the liver |
| | K77 | Liver disorders in disease classified elsewhere |
| **Asthma** | J45 | Asthma |
| **Non-haematological malignancy** | C0 | Malignant neoplasm of lip |
| | C1 | Malignant neoplasm of base of tongue |
| | C2 | Malignant neoplasm of other unspecified parts of tongue |
| | C3 | Malignant neoplasm of gum |
| | C4 | Malignant neoplasm of floor of mouth |
| | C5 | Malignant neoplasm of palate |
| | C6 | Malignant neoplasm of other and unspecified parts of mouth |
| | C7 | Malignant neoplasm of parotid gland |

| | | |
|---|---|---|
| **Haematological malignancy** | C8 | Malignant neoplasm of other and unspecified major salivary glands |
| | C9 | Malignant neoplasm of tonsil |
| **Stroke** | I63 | Cerebral infarction |
| | I65 | Occlusion and stenosis of precerebral arteries, not resulting in cerebral infarction |
| | I66 | Occlusion and stenosis of cerebral arteries, not resulting in cerebral infarction |
| **Chronic kidney disease** | N18.1-N18.5 | Chronic kidney disease stage 1-5 |
| | N18.9 | Chronic kidney disease, unspecified |
| | I13 | Hypertensive and renal disease |
| **Chronic heart disease** | I20 | Angina pectoris |
| | I21 | Acute myocardial infarction |
| | I22 | Subsequent myocardial infarction |
| | I23 | Certain current complications following acute myocardial infarction |
| | I24 | Other acute ischaemic heart diseases |
| | I25 | Chronic ischaemic heart disease |
| | I34 | Nonrheumatic mitral valve disorders |
| | I35 | Nonrheumatic aortic valve disorders |
| | I36 | Nonrheumatic tricuspid valve disorders |
| | I37 | Pulmonary valve disorders |
| | I42 | Cardiomyopathy |
| | I43 | Cardiomyopathy in diseases classified elsewhere |
| | I44 | Atrioventricular and left bundle-branch block |
| | I50 | Heart failure |
| **Immunocompromised** | D80 | Immunodeficiency with predominantly antibody defects |
| | D81 | Combined immunodeficiencies |
| | D82 | Immunodeficiency associated with other major defects |
| | D83 | Common variable immunodeficiency |
| | D84 | Other immunodeficiencies |
| **Dementia** | F01 | Vascular dementia |

| | | |
|---|---|---|
| | F02 | Dementia in other diseases classified elsewhere |
| | F03 | Unspecified dementia |
| | G30, G31 | Alzheimer disease & Other degenerative diseases of nervous system, not elsewhere classified |
| | F10.27 | Alcohol dependence, with alcohol-induced persisting dementia |
| | F10.97 | Alcohol use, unspecified with alcohol-induced persisting dementia |
| | F19.97 | Other psychoactive substance use, unspecified with psychoactive substance-induced persisting dementia |
| **Respiratory disease** | I27 | Other pulmonary heart diseases |
| | J6*-J7* | Lung diseases due to external agents |
| | J41 | Simple and mucopurulent chronic bronchitis |
| | J42 | Unspecified chronic bronchitis |
| | J43 | Emphysema |
| | J44 | Other chronic obstructive pulmonary disease |
| | J47 | Bronchiectasis |

**eTable 3:** Baseline cumulative subdistribution hazards for mortality in the final model, as needed for calculation of the 48 hour survival probabilities (eAppendix 5).

| **Time after landmark (hours)** | **Cumulative subdistribution hazard** | **Time after landmark (hours)** | **Cumulative subdistribution hazard** | **Time after landmark (hours)** | **Cumulative subdistribution hazard** | **Time after landmark (hours)** | **Cumulative subdistribution hazard** |
|---|---|---|---|---|---|---|---|
| 1 | 0.00094 | 13 | 0.00852 | 25 | 0.02143 | 37 | 0.03339 |
| 2 | 0.00161 | 14 | 0.00906 | 26 | 0.02339 | 38 | 0.03480 |
| 3 | 0.00255 | 15 | 0.00942 | 27 | 0.02463 | 39 | 0.03559 |
| 4 | 0.00304 | 16 | 0.00963 | 28 | 0.02525 | 40 | 0.03582 |
| 5 | 0.00363 | 17 | 0.01109 | 29 | 0.02673 | 41 | 0.03707 |
| 6 | 0.00427 | 18 | 0.01140 | 30 | 0.02783 | 42 | 0.03778 |
| 7 | 0.00458 | 19 | 0.01220 | 31 | 0.02876 | 43 | 0.03867 |
| 8 | 0.00516 | 20 | 0.01434 | 32 | 0.03008 | 44 | 0.04101 |
| 9 | 0.00563 | 21 | 0.01563 | 33 | 0.03095 | 45 | 0.04200 |
| 10 | 0.00713 | 22 | 0.01674 | 34 | 0.03182 | 46 | 0.04322 |
| 11 | 0.00787 | 23 | 0.01821 | 35 | 0.03265 | 47 | 0.04481 |
| 12 | 0.00834 | 24 | 0.01962 | 36 | 0.03318 | 48 | 0.04625 |

**eTable 4:** Final model coefficients for landmarks less than 28 days from admission or the first positive SARS-CoV-2 test, if infection was nosocomial.

| **Predictor** | **Coefficients when recorded** | **Coefficients if unrecorded**[1] |
|---|---|---|
| Age <75 years, at admission | -0.269 | – |
| Age <80 years, at admission | -0.135 | – |
| Heart rate, beats/min, mean during last 24h | 0.00154 | – |
| Respiratory rate, breaths/min, minimum during last 24h | 0.135 | – |
| SpO2/FiO2 ratio, minimum during last 24h | -0.0114 | – |
| WCC, $10^9$/L, most recent measurement during last 48h | 0.00169 | -0.000359 |
| Acidosis, 7.35 - (lowest pH during last 24h), or 0 if all above 7.35 | 5.97 | 1.66 |

**eTable 5**. Model coefficients for the alternative model with IL-6 replaced by CRP and re-calculating the model coefficients through the same penalised likelihood function used in the SCAD algorithm.

| **Predictor** | **Coefficients when recorded** | **Coefficients if unrecorded**[1] |
|---|---|---|
| Age <75 years, at admission | -0.115 | – |
| Age <80 years, at admission | -0.0582 | – |
| Clinical Frailty Score, at admission | 0.0672 | 0.150 |
| Heart rate, beats/min, mean during last 24h | 0.0128 | – |
| Respiratory rate, breaths/min, minimum during last 24h | 0.0515 | – |
| SpO2/FiO2 ratio, minimum during last 24h | -0.00346 | – |
| WCC, $10^9$/L, most recent measurement during last 48h | 0.00239 | -0.116 |
| Acidosis, 7.35 - (lowest pH during last 24h), or 0 if all above 7.35 | 2.73 | 0.474 |
| C-reactive protein, pg/ml, most recent measurement during last 48h | -0.0000350 | 0.220 |

1. If the predictor value is not recorded, the fixed value in this column is used, and the coefficient corresponding to the predictor value is ignored.

**eTable 6**: Baseline cumulative subdistribution hazards for the alternative model with IL-6 replaced by CRP, as needed for calculation of the 48 hour survival probabilities (eAppendix 5).

| Time after landmark (hours) | Cumulative subdistribution hazard | Time after landmark (hours) | Cumulative subdistribution hazard | Time after landmark (hours) | Cumulative subdistribution hazard | Time after landmark (hours) | Cumulative subdistribution hazard |
|---|---|---|---|---|---|---|---|
| 1 | 0.00010 | 13 | 0.00091 | 25 | 0.00224 | 37 | 0.00342 |
| 2 | 0.00017 | 14 | 0.00097 | 26 | 0.00243 | 38 | 0.00355 |
| 3 | 0.00028 | 15 | 0.00101 | 27 | 0.00255 | 39 | 0.00363 |
| 4 | 0.00033 | 16 | 0.00103 | 28 | 0.00262 | 40 | 0.00365 |
| 5 | 0.00039 | 17 | 0.00118 | 29 | 0.00276 | 41 | 0.00378 |
| 6 | 0.00046 | 18 | 0.00122 | 30 | 0.00287 | 42 | 0.00384 |
| 7 | 0.00049 | 19 | 0.00130 | 31 | 0.00296 | 43 | 0.00393 |
| 8 | 0.00056 | 20 | 0.00152 | 32 | 0.00309 | 44 | 0.00416 |
| 9 | 0.00061 | 21 | 0.00165 | 33 | 0.00318 | 45 | 0.00425 |
| 10 | 0.00077 | 22 | 0.00176 | 34 | 0.00326 | 46 | 0.00437 |
| 11 | 0.00084 | 23 | 0.00191 | 35 | 0.00334 | 47 | 0.00452 |
| 12 | 0.00089 | 24 | 0.00205 | 36 | 0.00340 | 48 | 0.00466 |

**eTable 7.** Final model coefficients for model omitting all blood tests.

| Predictor | Coefficients when recorded | Coefficients if unrecorded[1] |
|---|---|---|
| Age <75 years, at admission | -0.683 | – |
| Age <80 years, at admission | -0.195 | – |
| Clinical Frailty Score, at admission | 0.193 | -0.170 |
| Heart rate, beats/min, mean during last 24h | 0.0175 | – |
| Respiratory rate, breaths/min, minimum during last 24h | 0.0570 | – |
| SpO2/FiO2 ratio, minimum during last 24h | -0.0125 | – |

1. If the predictor value is not recorded, the fixed value in this column is used, and the coefficient corresponding to the predictor value is ignored.

**eTable 8**: Baseline cumulative subdistribution hazards for the alternative model without blood tests.

| Time after landmark (hours) | Cumulative subdistribution hazard | Time after landmark (hours) | Cumulative subdistribution hazard | Time after landmark (hours) | Cumulative subdistribution hazard | Time after landmark (hours) | Cumulative subdistribution hazard |
|---|---|---|---|---|---|---|---|
| 1 | 0.000687 | 13 | 0.006248 | 25 | 0.015666 | 37 | 0.024447 |
| 2 | 0.00118 | 14 | 0.006647 | 26 | 0.017095 | 38 | 0.02549 |
| 3 | 0.001872 | 15 | 0.006911 | 27 | 0.018004 | 39 | 0.026075 |
| 4 | 0.002229 | 16 | 0.007063 | 28 | 0.018459 | 40 | 0.026245 |
| 5 | 0.002662 | 17 | 0.008136 | 29 | 0.019544 | 41 | 0.027172 |
| 6 | 0.003129 | 18 | 0.008366 | 30 | 0.020346 | 42 | 0.027697 |
| 7 | 0.003357 | 19 | 0.008946 | 31 | 0.021037 | 43 | 0.028356 |
| 8 | 0.003784 | 20 | 0.010495 | 32 | 0.022007 | 44 | 0.030097 |
| 9 | 0.00413 | 21 | 0.011433 | 33 | 0.022648 | 45 | 0.030851 |
| 10 | 0.005232 | 22 | 0.012245 | 34 | 0.023292 | 46 | 0.031778 |
| 11 | 0.005773 | 23 | 0.013308 | 35 | 0.0239 | 47 | 0.032995 |
| 12 | 0.006119 | 24 | 0.014341 | 36 | 0.024293 | 48 | 0.034088 |

**eFigure 1.** Performance metrics for in-hospital mortality in the training dataset with 72 hour prediction horizon. (A) Receiver operator characteristic plot, with labels indicating the corresponding threshold and the dashed line indicating the line of no discrimination. (B) Precision-recall plot, with the 2.8% observed incidence indicated by the dashed line. (C) Number needed to evaluate against sensitivity. (D) Calibration plot (with 95% CI), by tenths of predicted risk and a LOESS interpolation (grey), with the dashed line indicating perfect calibration.

**eFigure 2.** Performance metrics for in-hospital mortality in the validation dataset with 72 hour prediction horizon. (A) Receiver operator characteristic plot, with labels indicating the corresponding threshold and the dashed line indicating the line of no discrimination. (B) Precision-recall plot, with the 3.1% observed incidence indicated by the dashed line. (C) Number needed to evaluate against sensitivity. (D) Calibration plot (with 95% CI), by tenths of predicted risk and a LOESS interpolation (grey), with the dashed line indicating perfect calibration.

**eTable 9.** Final model coefficients for 72 hour prediction horizon

| **Predictor** | **Coefficients when recorded** | **Coefficients if unrecorded**[1] |
|---|---|---|
| Age <75 years, at admission | -0.564 | – |
| Age <80 years, at admission | -0.183 | – |
| History of non-haematological malignancy | 0.146 | – |
| Clinical Frailty Score, at admission | 0.0721 | 0.0544 |
| Respiratory rate, breaths/min, minimum during last 24h | 0.100 | – |
| SpO2/FiO2 ratio, minimum during last 24h | -0.0111 | – |
| WCC, $10^9$/L, most recent measurement during last 48h | 0.00806 | -0.0198[a] |
| Acidosis, 7.35 - (lowest pH during last 24h), or 0 if all above 7.35 | 1.29 | 0.0.0898 |
| Platelets, $10^9$/L, most recent measurement during last 48h | -0.000375 | -0.0198[a] |

[a] WCC and Platelets are measured on the same sample, therefore the missingness for both clinical parameters is shared.

**eFigure 3.** Net Benefit for the dynamic model for threshold probabilities of 0-0.2 for patients of Wave 1.

**eFigure 4.** Net Benefit for the dynamic model for threshold probabilities of 0-0.2 for patients of Wave 2.

### Appendix

**eAppendix 1.** Diagnostic testing used either a real-time reverse transcription polymerase chain reaction (RT-PCR) of the RdRp gene from a nasopharyngeal swab, or the SAMBA II point-of-care test used at the hospital[e1]. Clinical diagnosis of COVID-19 was identified using International Classification of Diseases 10th Edition (ICD-10) codes in the EHR.

**eAppendix 2.**

These studies were the TACTIC-E and TACTIC-R trials (ISRCTN11188345 https://doi.org/10.1186/ISRCTN11188345), the REMAP-CAP platform trial for intensive care patients (ISRCTN67000769 https://doi.org/10.1186/ISRCTN67000769), and the RECOVERY trial (ISRCTN50189673 https://doi.org/10.1186/ISRCTN50189673).

**eAppendix 3.**

Patient consent was waived because the de-identified data presented here were collected during routine clinical practice; there was no requirement for informed consent.

**eAppendix 4.**

For patients for whom a CFS score had not been recorded by the treating team, a consultant or specialist registrar in Geriatric Medicine reviewed the clinical records and assigned a CFS score using only information recorded at the time of admission[32]. This approach has been shown to have good agreement with CFS scores assigned after face to face assessment (inter-rater reliability kappa 0.84)[33].

**eAppendix 5.**

Let $k_i \in \{0,\ldots,n_k\}$ be the event type of patient $i$, so that $T_{i,k}$ is patient $i$'s corresponding event time. The probability of patient $i$ with the clinical parameters (covariates) $X(s)$ at time s not experiencing any event before landmark s, and incurring event $k$ within time $t \in [0, w]$ of this landmark can therefore be calculated by:

$$\mathbb{P}(T_{i,k} < s + t \,|T_{i,k} \geq s, k_i = k, X(s)) = 1 - exp(-\int_{[0,\tilde{t}]} \lambda(u|X(s))du)$$

Here $\lambda(t|X(s))$is the sub-distribution hazard function for event $k$:

$$\lambda(t|X(s)) = \lambda_{k0}(t)\, exp(\beta^T X(s)),$$

with the subdistribution baseline hazard $\lambda_{k0}(t)$ for event $k$ at time $t \in [0, w]$. We report estimated values for β in Table 2. We report in eTable 3 the estimated cumulative baseline sub-distribution hazard function $\int_{[0,t]} \lambda_{k0}(u)\, du$. For the model using CRP in place of IL-6, see eTable 4 and eTable 5.

To derive the 48-hour mortality probability $(k_i = k_{mort},\ w = 48h)$ from the Fine-Gray model, we therefore compute the probability of the mortality occurring between the landmarking time *s* and the end of the prediction horizon *s*+*w*, given the patient did not have any event prior to the landmark and accounting for the covariates $X(s)$ collected at the landmark time $s$.

$$\mathbb{P}(T_{i,k} < s + w | T_{i,k} \geq s, k_i = k_{mort}, X(s)) = 1 - exp(- \int_{[0,w]} \lambda(t|X(s))dt)$$

To allow for easier access a web-app is available on http://shiny.mrc-bsu.cam.ac.uk/apps/covid19mortalityrisk/, that calculates the 48-hour mortality probability based on user input.
